# The impact of mental health and substance use issues on COVID-19 vaccine readiness: a cross sectional community-based survey in Ontario, Canada

**DOI:** 10.1101/2021.08.30.21262844

**Authors:** Kamna Mehra, Roula Markoulakis, Sugy Kodeeswaran, Donald A. Redelmeier, Mark Sinyor, James MacKillop, Amy Cheung, Emily E. Levitt, Tracey Addison, Anthony Levitt

## Abstract

**Background:** COVID-19 vaccines have been approved for use in Canada since December 2020. However, data about factors associated with vaccine hesitancy and the impact of mental health and/or substance use (MHSU) issues on vaccine uptake are currently not available. The goal of this study was to explore factors, particularly MHSU factors, that impact COVID-19 vaccination intentions in Ontario, Canada.

**Methods:** A community-based cross-sectional survey with recruitment based on age, gender, and geographical location (to ensure a representative population of Ontario), was conducted in February 2021. Multinomial logistic regression was used to test the relationship between COVID-19 vaccination status and plans and sociodemographic background, social support, anxiety about contracting COVID-19, and MHSU concerns.

**Results:** Of the total sample of 2528 respondents, 1932 (76.4%) were vaccine ready, 381 (15.1%) were hesitant, and 181 (7.1%) were resistant. Significant independent predictors of vaccine hesitancy compared with vaccine readiness included younger age (OR=2.11, 95%CI=1.62-2.74), female gender (OR=1.36, 95%CI=1.06-1.74), Black ethnicity (OR=2.11, 95%CI=1.19-3.75), lower education (OR=1.69, 95%CI=1.30-2.20), lower SES status (OR=.88, 95%CI=.84-.93), lower anxiety about self or someone close contracting COVID-19 (OR=2.06, 95%CI=1.50-2.82), and lower depression score (OR=.90, 95%CI=.82-.98). Significant independent predictors of vaccine resistance compared with readiness included younger age (OR=1.72, 95%CI=1.19-2.50), female gender (OR=1.57, 95%CI=1.10-2.24), being married (OR=1.50, 95%CI=1.04-2.16), lower SES (OR=.80, 95%CI=.74-.86), lower satisfaction with social support (OR=.78, 95%CI=.70-.88), lower anxiety about contracting COVID-19 (OR=7.51, 95%CI=5.18-10.91), and lower depression score (OR=.85, 95%CI=.76-.96).

**Interpretation:** COVID-19 vaccination intention is affected by sociodemographic factors, anxiety about contracting COVID-19, and select mental health issues.

## Introduction

Identifying predictors of COVID-19 vaccination hesitancy and resistance is crucial to implementing a successful vaccination campaign. In April 2020, a study conducted in the United States by Fisher et al.^1^ found younger age, Black race, lower education, and prior missed vaccinations were independent predictors of COVID-19 vaccination hesitancy. Similarly, a study conducted in the United Kingdom and Ireland found that age, gender, ethnicity, geographical location, socioeconomic status (SES), and political affiliation were associated with COVID-19 vaccination hesitancy.^2^ A recent survey in Ontario, Canada,^3^ found 17.2% participants were unwilling to receive the COVID-19 vaccine, and that females and those with lower education were more likely to be unwilling.; however, data on the impact of MHSU issues on COVID-19 vaccination acceptance in Canada are currently unavailable.^7^ Compared with the general population, people living with mental health and substance use (MHSU) issues have been shown to be at higher risk of contracting, and to have higher morbidity and mortality from, COVID-19 infection.^4,5^ Adverse medical outcomes from contracting COVID-19 are more substantial for people with severe mental illness than for those with less severe mental illness.^6^ Thus, the goal of the current study was to understand the predictors of COVID-19 vaccination hesitancy and resistance in a representative population of the province of Ontario, Canada, with a focus on the impact of MHSU issues on these intentions.

## Methods

### Participants and Procedures

The current study involved a community-based, cross-sectional survey of 2528 Ontarians, age 18 years or older, recruited as a provincially representative sample through a respondent panel: Delvinia’s AskingCanadians. Individuals registered with AskingCanadians were randomly sent a unique survey link. Interlocking quotas, based on known age, gender, and regional population (Toronto, Southwestern, Eastern, Central, and Northern) proportions were used.^6,7^ The aim of the interlocking quotas for age and gender was to ensure provincial representativeness based on a difference less than five percentage points or within five percent of the mean. Although regional population quotas were utilized, lower density regions outside of the largest city, Toronto, were oversampled to ensure adequate sample across all five regions of Ontario, based on the power needed to detect the known prevalence of depression in each region, within a 5% margin of error.^8^

The study was approved by the Sunnybrook Health Sciences Centre Research Ethics Board. Participants gave informed consent prior to participating. The survey comprised 63 items and was open from February 22 to March 15, 2021. Of those participants who were eligible and provided consent, the survey completion rate was 79%.

### Measures

#### COVID-19 vaccination

Participants were asked about COVID-19 vaccination intention with responses reflecting whether or not they planned to get COVID-19 vaccination once available/eligible, had received the first or both doses of the vaccination, had heard about the vaccination but were undecided, had heard about it and did not plan to be vaccinated, and had not heard about the vaccination.

#### Sociodemographic data and COVID-19 related information

Age, gender, ethnicity, geographical location, education level, marital status, SES,^9^ and living situation information was collected. Satisfaction with social support since the onset of the pandemic (e.g., friends, family, community, co-workers, pets, etc.) was assessed via seven-point bipolar Likert scale (1=extremely satisfied/7=extremely dissatisfied). COVID-19 exposure risk was assessed using an original 12-item checklist, with responses reflecting higher risk (being tested and diagnosed with COVID-19, being told they had COVID-19 by a professional, someone they lived with diagnosed with COVID-19, someone else close to them diagnosed with COVID-19, someone close to them passed away as a result of COVID-19) and lower risk (suspecting they had COVID-19 but not diagnosed, tested for COVID-19 but negative, someone they lived with was tested for COVID-19, someone close to them was tested for COVID-19, self-quarantine due to travel, healthcare worker dealing with COVID-19 patients, confirmed COVID-19 cases at places they visit). Fear of self and/or someone close contracting COVID-19 was assessed on two separate items (but later grouped together for analysis) via five-point Likert scale (1=extremely worried/5=not worried at all).

#### MHSU concerns

The presence of mental health concerns was assessed by the American Psychiatric Association’s Diagnostic Statistical Manual (DSM)-5 Self-Rated Level 1 Cross-Cutting Symptom Measure (CCSM), Adult version.^10^ This measure has good test-retest reliability and is clinically useful in Canadian samples.^11^ The Cronbach’s alpha, in the current study, for subscales with more than 1 item ranged from .69 to .85 and for the overall scale (20 items) was 0.94. Substance use was assessed by the World Health Organization Alcohol, Smoking, and Substance Involvement Screening Test (ASSIST v3.0).^12^ Substances assessed included tobacco, alcohol, cannabis, and opioids. This scale categorizes participants into low, moderate, and high risk categories for substance use disorder and has good to excellent internal validity.^13^

### Statistical Analysis

Statistical analysis was conducted using SPSS 26.0. Vaccination intention was divided into those who: planned to receive the vaccination or had received the first/both doses (vaccine ready), were undecided (vaccine hesitant), or did not plan to receive the vaccination (vaccine resistant). Univariate analyses were conducted using chi-square tests and ANOVA to compare vaccine ready, hesitant, and resistant based on sociodemographic variables, COVID-19-related variables, and MHSU variables. Multinomial logistic regression was then conducted to compare vaccine hesitant or resistant to vaccine ready participants. Variables included in the logistic regression were those that were significant in the univariate analyses. In the multinomial logistic regression which compared vaccine ready participants with both vaccine hesitant or resistant participants, the independent variables included were age (median split, since continuous variable did not have a linear relationship with its logit), gender (women vs. men), ethnicity (Non-Caucasian vs. Caucasian), geographical location (GTA vs. outside GTA), education (completed college/university vs. not completed college/university), marital status (married vs. unmarried), and anxiety about self and/or someone close contracting COVID-19 (not worried vs. worried, since the continuous variable did not have a linear relationship with its logit). Scores on the depression, anxiety, suicidal ideation, psychosis, and repetitive thoughts and behaviors sub-scales from the DSM-5 CCSM, social support, and SES since onset of pandemic were entered as continuous variables. Tobacco, cannabis, and opioid use were entered as categorical variables (no/low risk vs moderate/high risk).

## Results

The mean age of the study sample was 48.26 years (within 5% of the provincial average age of adults 18 years or older of 48.14 years.)^14^ The gender distribution was 49.8% women and 48.9% men (within 5% of the provincial distribution of 51.2% women and 48.8% men.)^15^

Out of 2528 participants, 1819 (72.0%) planned to get COVID-19 vaccination when available to them, 58 (2.3%) had received the first dose of the vaccination, 55 (2.2%) had received both the first and second dose of the vaccination, 381 (15.1%) had heard about the vaccination but were undecided, 181 (7.2%) had heard about it and did not plan to be vaccinated, and 34 (1.3%) had not heard about the vaccination.

### Univariate tests for vaccination intention: Sociodemographic variables

Based on chi-square analysis, there was significant difference between vaccine ready, vaccine hesitant, and vaccine resistant participants based on sociodemographic variables (Table 1). Those who were younger than the median age of 47 years (χ^2^=50.751, p<.001), women (χ^2^=5.378, p=.020), unmarried (χ^2^=6.173, p=.013), or had not completed college/university (χ^2^=24.243, p<.001) were significantly more likely to be vaccine hesitant than vaccine ready. Those who were younger (χ^2^= 16.826, p<.001), living outside the Greater Toronto Area (GTA) (χ^2^= 6.078, p=.014), or had not completed college/university (χ^2^=10.665, p=.001) were significantly more likely to be vaccine resistant than vaccine ready. The ANOVA comparing SES since the onset of the pandemic across those who were vaccine ready (M=6.78, SD=2.269), vaccine hesitant (M=5.90, SD=2.381), and vaccine resistant (M=5.31, SD=2.682), was significant (F(49.888), df(2), p<.001). Post hoc tests, using LSD, revealed participants with lower SES were more likely to be vaccine hesitant (MD=.88, p<.001) and vaccine resistant (MD=1.46, p<.001) than vaccine ready.

**Table 1:**
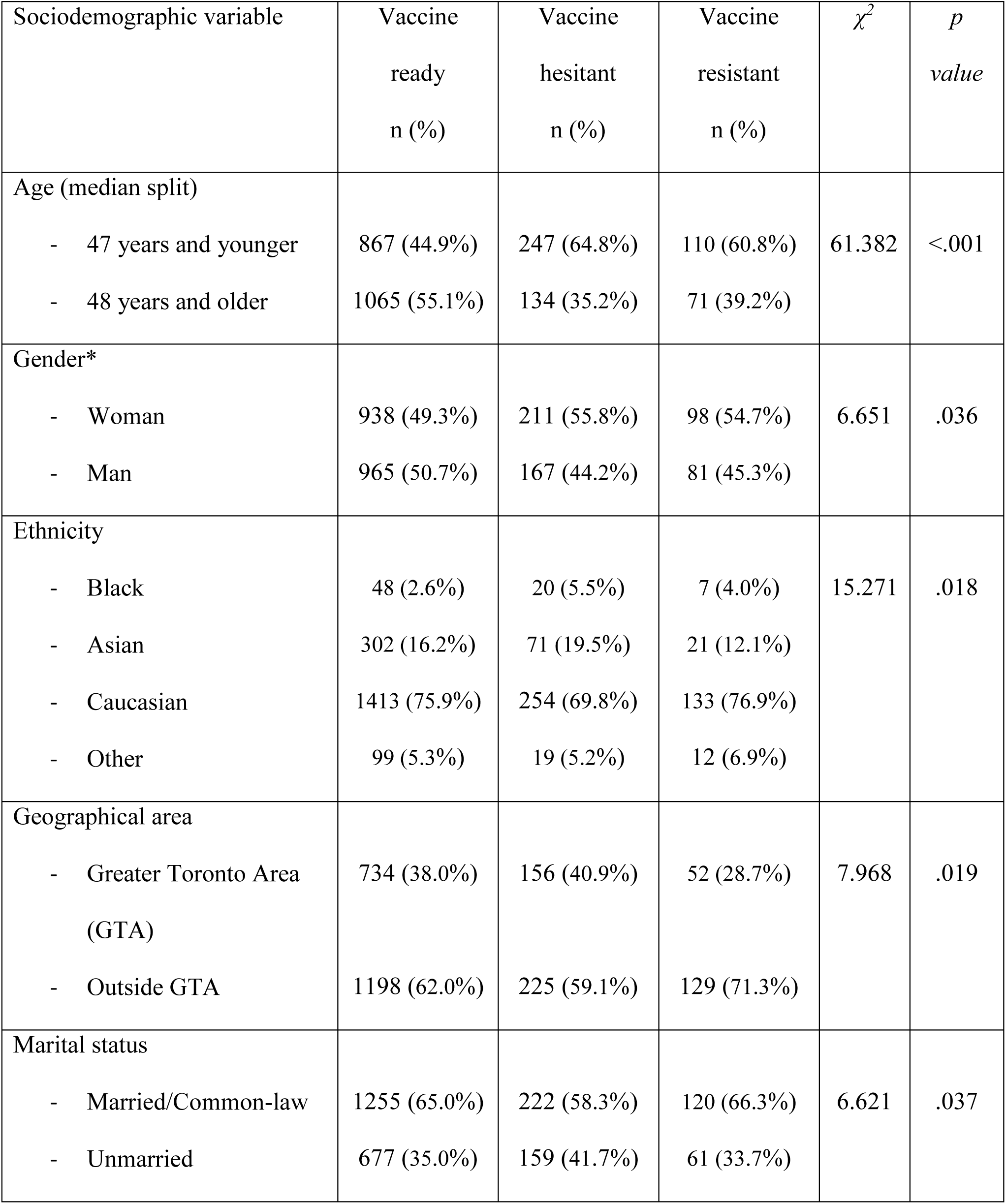
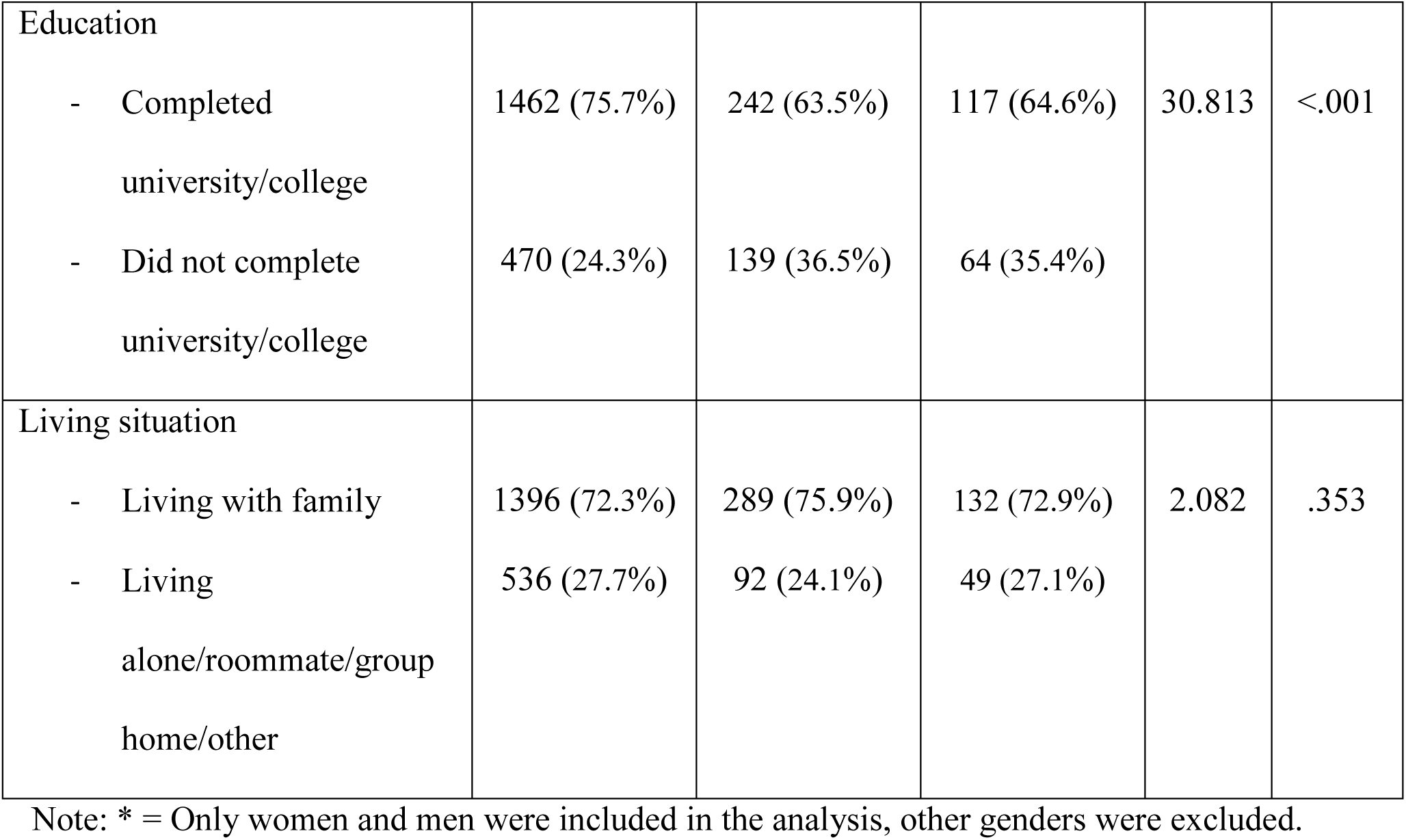
Sociodemographic information of survey respondents, based on the likelihood of getting COVID-19 vaccination.

### Univariate tests for vaccination intention: COVID-19 related variables

Using ANOVA, there was a significant difference in mean social support since pandemic onset across those who were vaccine ready (M=4.52, SD=1.502), vaccine hesitant (M=4.33, SD=1.487), and vaccine resistant (M=3.88, SD=1.618; F(16.440), df(2), p<.001). Post hoc testing (LSD) revealed that vaccine hesitant (MD=.194, p=.022) and vaccine resistant (MD= .644, p<.001) participants were less satisfied with social support as compared with vaccine ready participants. Those who were vaccine hesitant (χ^2^=17.376, p<.001) and vaccine resistant (χ^2^=176.384, p<.001) were significantly more likely to be worried about self and/or someone close contracting COVID-19 (Table 2). There was no significant difference between vaccine hesitancy, resistance, and readiness based on risk of COVID-19 (Table 2).

**Table 2:** COVID-19 related variables and likelihood of getting COVID-19 vaccination.

| COVID-19 related variable | Vaccine ready<br>n (%) | Vaccine hesitant<br>n (%) | Vaccine resistant<br>n (%) | $\chi^2$ | *p value* |
| --- | --- | --- | --- | --- | --- |
| COVID-19 anxiety self or someone close |  |  |  |  |  |
| - Worried | 1704 (88.2%) | 306 (80.3%) | 93 (51.4%) | 175.159 | <.001 |
| - Not worried | 228 (11.8%) | 75 (19.7%) | 88 (48.6%) |  |  |
| COVID-19 risk |  |  |  |  |  |
| - High risk | 323 (16.7%) | 71 (18.6%) | 23 (12.7%) | 3.097 | .213 |
| - Low risk | 1609 (83.3%) | 310 (81.4%) | 158 (87.3%) |  |  |

### Univariate tests for vaccination intention: Mental health issues

Using ANOVA, there were significant differences between the mean scores on depression, suicidal ideation, anxiety, psychosis, and repetitive thoughts and behaviors sub-scales based on vaccination intention (Table 3). Based on post-hoc LSD analysis, higher mean scores in depression (MD=-.25, p=0.040), anxiety (MD=-.61, p<.001), suicidal ideation (MD=-.16, p<.001), psychosis (MD=-.22, p=0.001), and repetitive thoughts and behaviors (MD=-.39, p<.001) were associated with vaccine hesitancy rather than vaccine readiness. Higher mean scores for anxiety (MD=-.48, p=.044), suicidal ideation (MD=-.20, p<.001), psychosis (MD=-.38, p<.001), and repetitive thoughts and behaviors (MD=-.51, p<.001) were also significantly associated with vaccine resistance rather than vaccine readiness.

**Table 3:** Mental health issues and likelihood of getting COVID-19 vaccination.

| Mental health issues | Vaccine<br>ready<br>M (SD) | Vaccine<br>hesitant<br>M (SD) | Vaccine<br>resistant<br>M (SD) | F | df | *p value* |
| --- | --- | --- | --- | --- | --- | --- |
| Depression | 2.30 (2.11) | 2.55 (2.23) | 2.58 (2.27) | 3.149 | 2 | .043 |
| Suicidal ideation | .24 (.68) | .40(.85) | .44 (.90) | 12.317 | 2 | <.001 |
| Anxiety | 3.00 (3.01) | 3.61 (3.15) | 3.48 (3.33) | 7.594 | 2 | .001 |
| Psychosis | .38 (1.12) | .60 (1.39) | .76 (1.65) | 12.209 | 2 | <.001 |
| Repetitive thoughts<br>and behaviors | 1.21 (1.71) | 1.60 (1.92) | 1.72 (2.04) | 12.973 | 2 | <.001 |

### Univariate tests for vaccination intention: Substance use issues

There was a significant difference between vaccine ready, hesitant, and resistant participants in the proportion with moderate/high risk for substance use disorder (Table 4). Those who were vaccine hesitant were significantly more likely to be associated with moderate/high risk of tobacco use disorder (χ^2^=6.513, p=.011), cannabis use disorder (χ^2^=6.679, p=.010), and opioid use disorder (χ^2^=4.871, p=.027) compared to vaccine ready; and vaccine resistance was significantly more likely to be associated with moderate/high risk of tobacco use disorder (χ^2^= 8.906, p=.003), cannabis use disorder (χ^2^= 9.926, p=.002), and opioid use disorder (χ^2^= 13.532, p<.001) compared to vaccine readiness.

**Table 4:**
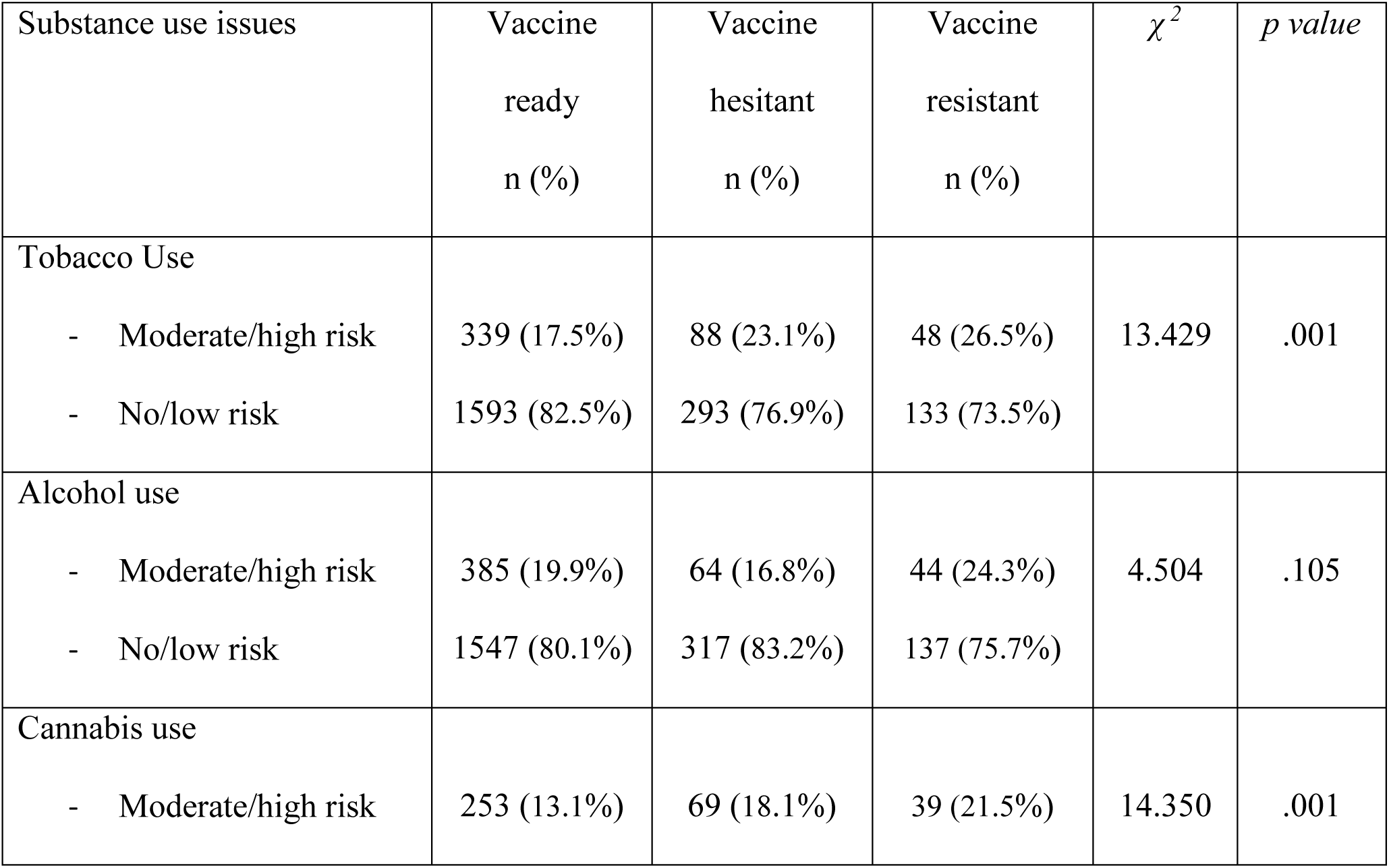
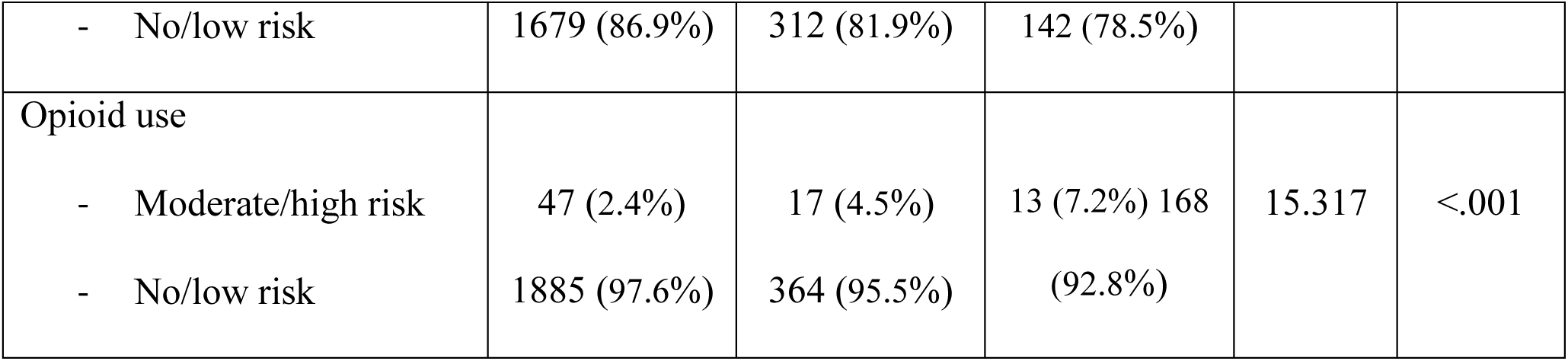
Substance use issues and likelihood of getting COVID-19 vaccination.

### Multinomial logistic regression tests for vaccination intention

Multinomial logistic regression modelling (Table 5) revealed variables that independently and significantly predicted vaccine hesitancy rather than vaccine readiness were: younger age (OR=2.11, 95%CI=1.62-2.74), female gender (OR=1.36, 95%CI=1.06-1.74), Black ethnicity (OR=2.11, 95%CI=1.19-3.75), lower education (OR=1.69, 95%CI=1.30-2.19), lower SES (OR=.88, 95%CI=.84-.93), lower anxiety about contracting COVID-19 (OR=2.06, 95%CI=1.50-2.82), and lower depression score (OR=.90, 95%CI=.82-.98). Significant independent predictors of vaccine resistance rather than vaccine readiness included younger age (OR=1.72, 95%CI=1.19-2.50), female gender (OR=1.57, 95%CI=1.10-2.24), being married (OR=1.50, 95%CI=1.04-2.16), lower SES (OR=.80, 95%CI=.74-.86), lower satisfaction with social support (OR=.78, 95%CI=.70-.88), lower anxiety about contracting COVID-19 (OR=7.51, 95%CI=5.18-10.91), and lower depression score (OR=.85, 95%CI=.76-.96).

**Table 5:** Multinomial logistic regression comparing vaccine ready to vaccine hesitant and vaccine resistant.

| Variables | Predictor | Reference | Vaccine hesitant vs. Vaccine ready |  |  | Vaccine resistant vs. Vaccine ready |  |  |
| --- | --- | --- | --- | --- | --- | --- | --- | --- |
|  |  |  | p | OR | 95% CI | p | OR | 95% CI |
| **Categorical variables** |  |  |  |  |  |  |  |  |
| Age | 47 years or younger | 48 years or older | <.001* | 2.11 | 1.62-2.74 | .004* | 1.72 | 1.19-2.50 |
| Gender | Female | Male | .014* | 1.36 | 1.06-1.74 | .013* | 1.57 | 1.10-2.24 |
| Ethnicity | Black | Caucasian | .011* | 2.11 | 1.19-3.75 | .081 | 2.19 | .91-5.28 |
|  | Asian |  | .548 | 1.11 | .79-1.56 | .642 | .88 | .50-1.53 |
|  | Other |  | .683 | .89 | .52-1.53 | .932 | .97 | .46-2.03 |
| Geographical location | Outside GTA | GTA | .407 | .90 | .69-1.16 | .187 | 1.31 | .88-1.96 |
| Education | Did not complete college/university | Completed college/university | <.001* | 1.69 | 1.30-2.20 | .055 | 1.44 | .99-2.08 |
| Marital status | Married | Unmarried | .602 | .94 | .73-1.20 | .031* | 1.50 | 1.04-2.16 |
| Worry about contracting COVID-19 | Not worried | Worried | <.001* | 2.06 | 1.50-2.82 | <.001* | 7.51 | 5.18-10.91 |
| Tobacco use | No/low risk | Moderate/high risk | .167 | .81 | .60-1.09 | .230 | .77 | .50-1.18 |
| Cannabis use | No/low risk | Moderate/high risk | .451 | .88 | .62-1.24 | .150 | .71 | .45-1.13 |
| Opioid use | No/low risk | Moderate/high risk | .192 | .66 | .35-1.24 | .303 | .66 | .30-1.46 |
| **Continuous variables** |  |  |  |  |  |  |  |  |
| SES since pandemic |  |  | <.001* | .88 | .84-.93 | <.001* | .80 | .74-.86 |
| Social support since pandemic |  |  | .182 | .95 | .87-1.03 | <.001* | .78 | .70-.88 |
| Depression |  |  | .013* | .90 | .82-.98 | .010* | .85 | .76-.96 |
| Anxiety |  |  | .959 | 1.002 | .94-1.07 | .614 | .98 | .89-1.07 |
| Suicidal ideation |  |  | .130 | 1.17 | .96-1.42 | .077 | 1.28 | .97-1.68 |
| Psychosis |  |  | .664 | .97 | .87-1.09 | .285 | 1.09 | .93-1.28 |
| Repetitive thoughts and behaviors |  |  | .112 | 1.09 | .98-1.20 | .206 | 1.10 | .95-1.27 |

Note: \* = Significance at the .05 level, Full model $\chi^2$ (38) = 352.556, p < .001, Nagelkerke
R<sup>2</sup>=.186

### Interpretation

This study found that higher scores on the depression measure independently differentiated participants who were vaccine ready compared to those who were hesitant or resistant; a novel finding, to the best of our knowledge. Depressed participants may be more introspective or concerned with their health, leading to higher readiness for vaccination. Alternatively, participants with depression may see vaccination as a concrete way to remove stressors from their lives. In addition, depressed participants may experience cognitive distortions contributing to negative interpretations of the pandemic,^16^ contributing to vaccine readiness. A study conducted by Palgi et al.^17^ found that among those who had already received the first dose of the vaccination, vaccine hesitancy was related to higher levels of depression. Our results do not support that finding, which may reflect a difference in the sampling, the tools used to assess mental health issues, or geographic or cultural differences. Additionally, in the current study, no other MHSU issues, including anxiety disorders, significantly differentiated those who were vaccine hesitant or resistant from those who were vaccine ready. This finding suggests it is not reasonable to ascribe vaccine hesitancy or resistance to any large degree to most MHSU issues. However, depression was found to be associated with vaccine readiness and this relationship warrants further exploration.

Age, gender, ethnicity, education, marital status, and SES were found to be significant independent predictors of vaccine hesitancy or resistance, with younger participants, females, and those who identified with lower SES since the onset of the pandemic significantly more likely to be vaccine hesitant and resistant, similar to previous studies in Canada and other countries.^1–3,18^ Lower level of education and identifying with Black ethnicity significantly distinguished between vaccine readiness and vaccine hesitancy, but not between vaccine readiness and resistance. Of note, marital status was the only other variable that distinguished the hesitant and resistant participants, such that unmarried participants were less likely to be vaccine resistant. Exploring the possible reasons for being undecided but not unwilling are out of the scope of the current study and have been explored in other studies.^2^ Nonetheless, this finding might suggest that more intensive education and appropriate information targeting these groups may enhance the proportion of the population who might be vaccine ready in the future.

Those participants who were vaccine resistant were significantly less satisfied with their social support since the onset of the pandemic than those who were vaccine ready. It is possible that people with fewer social interactions and lower satisfaction with those interactions may have less communication about COVID-19 vaccination through friends, family members, co-workers, community groups, etc. and therefore less information about the potential benefits of vaccination. Furthermore, people who are happy with their social interactions may have the desire to continue or to expand their interactions, leading them to be more receptive to vaccination. Similar to previous findings, participants who were vaccine ready were more worried about themselves or someone close to them contracting COVID-19.^18^ Thus, consistent with other evidence, relationships may play a crucial role in COVID-19 vaccination intention, in that those with close relationships may feel a sense of social duty or moral obligation to protect those around them,^19^ or the fear of contracting the COVID-19 infection may lend to vaccine readiness.^18,20^

### Limitations

This study was limited to Ontario, and may not be generalizable outside the province. Furthermore, the sample was randomly selected from participants registered with a respondent panel, and the survey was only available online and in the English language. Although this group may not be representative of the entire population of Ontario, the sample had similar characteristics to the population of Ontario.^14,15^ Moreover, the vaccine readiness of individuals of a small proportion of the population may change over time.^21^ This cross-sectional design was also only able to identify associations between variables, and did not allow us to determine whether these findings have been stable as the vaccine rollout has progressed. For example, the spread of the COVID-19 Delta variant, after the current survey was conducted, may affect vaccine readiness. While other studies have shown stability in vaccine readiness over time,^22,23^ study of factors influencing vaccine readiness over time is warranted. Furthermore, the DSM-5 CCSM has been validated for use as a screening questionnaire, and is not meant to reflect severity or diagnosis of mental illness. Nonetheless, the internal reliability of the severity measure was adequate based on the excellent Cronbach’s alpha.

## Conclusion

This study provides data to decision makers to take into account as they work to enhance vaccine uptake and to ensure that certain populations are provided targeted and helpful information to make an informed choice about COVID-19 vaccination.

## Data Availability

Data presented in this manuscript is available upon reasonable request.

## Data access

Data presented in this manuscript is available upon reasonable request.

## Acknowledgements

None

## Conflict of interest

None

## Funding

This study was funded by the Sunnybrook Foundation & Sunnybrook Research Institute, COVID-19 Research Initiative. MS is supported by Academic Scholar Awards from the Departments of Psychiatry at Sunnybrook Health Sciences Centre and the University of Toronto.

## Supplemental material

None

## Notes

### Competing Interest Statement

The authors have declared no competing interest.

### Author Declarations

The study was approved by the Sunnybrook Health Sciences Centre Research Ethics Board.

